## Supplementary Materials for "Immune boosting and the perils of interpreting pertussis seroprevalence studies"

Matthieu Domenech de Cellès[1,*], Anabelle Wong[1,2,*], Tine Dalby[3], Pejman Rohani[4,5,6]

1. Max Planck Institute for Infection Biology, Infectious Disease Epidemiology group, Chariteplatz 1, 10117 Berlin, Germany
2. Institute of Public Health, Charité – Universitätsmedizin Berlin, Charitéplatz 1, 10117 Berlin, Germany
3. Department of Infectious Disease Epidemiology and Prevention, Statens Serum Institut, 2300 Copenhagen S, Denmark
4. Odum School of Ecology, University of Georgia, Athens, Georgia, USA
5. Center of Ecology of Infectious Diseases, Athens, Georgia, USA
6. Department of Infectious Diseases, College for Veterinary Medicine, University of Georgia, Athens, Georgia, USA

Corresponding author: Dr. Matthieu Domenech de Cellès. Address: Max Planck Institute for Infection Biology, Charitéplatz 1, Campus Charité Mitte, 10117 Berlin, Germany.

*These authors contributed equally.

### Supplementary Methods

#### *Review of pertussis seroprevalence studies*

We aimed to review recent seroprevalence studies conducted on the general population to understand how seroprevalence data were interpreted and whether immune boosting was discussed. Nineteen studies were included. We detail the literature search and study selection procedures below.

Following the search terms used by a previous review by Barkoff et al. (1), we conducted a search for articles published in the past five years (as of the search date, February 3, 2025) that contain [pertussis AND seroprevalence], [pertussis AND serosurvey], [pertussis AND serosurveillance] OR [pertussis AND seroincidence] in the Title/Abstract on the literature databased, PubMed, and found 59 articles.

Upon initial screening, 11 records were excluded due to article type or language. The remaining records underwent the abstract retrieval and screening stage, during which 21 records were excluded due to (i) study type (assay method development); (ii) study aim (investigating antibody waning among vaccinated children or examining factors associated with antibody concentrations instead of assessing seroprevalence in the sample); or (iii) being conducted in specific risk groups (pregnant women, healthcare workers [HCW], or patients with chronic obstructive pulmonary disease [COPD]).

Full-text articles of the remaining records were retrieved and assessed for eligibility, resulting in an additional eight studies being excluded due to (i) article type, (ii) study type (simulations or modeling), (iii) study aim (antibody waning among vaccinated children or factors associated with antibody concentrations), or (iv) being conducted in specific risk groups (adults of child-bearing age).

***Model formulation***

Force of infection

The force of infection in age group *i* was expressed as:

$$\lambda_i = q_i \sum_j M_{ij} \frac{I_1^{(j)} + \theta I_2^{(j)} + \iota}{N_j}$$

Here, $q_i$ represents the age-specific susceptibility to infection, fixed to 0.09 in individuals aged 0–9 yrs, 0.05 in 10–19 yrs, and 0.008 in ≥20 yrs, based on previous fits in Massachusetts, USA (2). The relative transmissibility of secondary infections was fixed based on the same source ($\theta = 0.99$). The parameters $M_{ij}$ represent the contact rates between age groups *i* and *j* and form the social contact matrix (SCM) *M*, defined on the intensive scale (as per (3)). These SCMs were fixed based on the work of Mistry *et al.* (4). The parameter ι represents the prevalence of imported cases, set to a small value ($\iota = 10^{-3}$) to prevent stochastic extinctions. The age-specific population sizes $N_j$ were fixed based on 2010 demographic data in every country, available from Mistry *et al.* (4).

Deterministic equations

The model variables are listed in Table S3. The deterministic variant of the model was described by a set of ordinary differential equations, given below for newborns (age group $i = 0$, the superscript indicates the age group):

$$
\begin{aligned}
\frac{dS_1^{(0)}}{dt} &= bN - (\lambda_0 + \delta_0)S_1^{(0)} \\
\frac{dE_1^{(0)}}{dt} &= \lambda_0 S_1^{(0)} - (\sigma + \delta_0)E_1^{(0)} \\
\frac{dI_1^{(0)}}{dt} &= \sigma E_1^{(0)} - (\gamma + \delta_0)I_1^{(0)} \\
\frac{dS_2^{(0)}}{dt} &= \alpha_R R^{(0)} + \alpha_V V^{(0)} - (\lambda_0 + \delta_0)S_2^{(0)} \\
\frac{dE_2^{(0)}}{dt} &= \lambda_0 S_2^{(0)} - (\sigma + \delta_0)E_2^{(0)} \\
\frac{dI_2^{(0)}}{dt} &= \sigma E_2^{(0)} - (\gamma + \delta_0)I_2^{(0)} \\
\frac{dR_{P,1}^{(0)}}{dt} &= \gamma(I_1^{(0)} + I_2^{(0)}) - (1/t_n + \delta_0)R_{P,1}^{(0)} \\
\frac{dR^{(0)}}{dt} &= R_{P,1}^{(0)}/t_n + R_{P,2}^{(0)}/t_n - (\alpha_R + \rho_R\lambda_0 + \delta_0)R^{(0)} \\
\frac{dR_E^{(0)}}{dt} &= \rho_R\lambda_0 R^{(0)} - (1/t_p + \delta_0)R_E^{(0)} \\
\frac{dR_{P,2}^{(0)}}{dt} &= R_E^{(0)}/t_p - (1/t_n + \delta_0)R_{P,2}^{(0)} \\
\frac{dV^{(0)}}{dt} &= 0 \\
\frac{dV_E^{(0)}}{dt} &= 0 \\
\frac{dV_P^{(0)}}{dt} &= 0
\end{aligned}
$$

In the older age groups ($i \geq 1$), the dynamic was governed by the following set of ODEs:

$$\begin{aligned}
\frac{dS_1^{(i)}}{dt} &= \delta_{i-1}S_1^{(i-1)} - (\lambda_i + v_i + \delta_i)S_1^{(i)} \\
\frac{dE_1^{(i)}}{dt} &= \delta_{i-1}E_1^{(i-1)} + \lambda_i S_1^{(i)} - (\sigma + \delta_i)E_1^{(i)} \\
\frac{dI_1^{(i)}}{dt} &= \delta_{i-1}I_1^{(i-1)} + \sigma E_1^{(i)} - (\gamma + \delta_i)I_1^{(i)} \\
\frac{dS_2^{(i)}}{dt} &= \delta_{i-1}S_2^{(i-1)} + \alpha_R R^{(i)} + \alpha_V V^{(i)} - (\lambda_i + v_i + \delta_i)S_2^{(i)} \\
\frac{dE_2^{(i)}}{dt} &= \delta_{i-1}E_2^{(i-1)} + \lambda_i S_2^{(i)} - (\sigma + \delta_i)E_2^{(i)} \\
\frac{dI_2^{(i)}}{dt} &= \delta_{i-1}I_2^{(i-1)} + \sigma E_2^{(i)} - (\gamma + \delta_i)I_2^{(i)} \\
\frac{dR_{P,1}^{(i)}}{dt} &= \delta_{i-1}R_{P,1}^{(i-1)} + \gamma(I_1^{(i)} + I_2^{(i)}) - (1/t_n + \delta_i)R_{P,1}^{(i)} \\
\frac{dR^{(i)}}{dt} &= \delta_{i-1}R^{(i-1)} + R_{P,1}^{(i)}/t_n + R_{P,2}^{(i)}/t_n - (\alpha_R + \rho_R\lambda_i + \delta_i)R^{(i)} \\
\frac{dR_E^{(i)}}{dt} &= \delta_{i-1}R_E^{(i-1)} + \rho_R\lambda_i R^{(i)} - (1/t_p + \delta_i)R_E^{(i)} \\
\frac{dR_{P,2}^{(i)}}{dt} &= \delta_{i-1}R_{P,2}^{(i-1)} + R_E^{(i)}/t_p - (1/t_n + \delta_i)R_{P,2}^{(i)} \\
\frac{dV^{(i)}}{dt} &= \delta_{i-1}V^{(i-1)} + v_i(S_1^{(i)} + S_2^{(i)}) + V_P^{(i)}/t_n - (\alpha_V + \rho_V\lambda_i + \delta_i)V^{(i)} \\
\frac{dV_E^{(i)}}{dt} &= \delta_{i-1}V_E^{(i-1)} + \rho_V\lambda_i V^{(i)} - (1/t_p + \delta_i)V_E^{(i)} \\
\frac{dV_P^{(i)}}{dt} &= \delta_{i-1}V_P^{(i-1)} + V_E^{(i)}/t_p - (1/t_n + \delta_i)V_P^{(i)}
\end{aligned}$$

Here, $b$ represents the per capita birth rate and $N$ the total population size, two parameters we fixed based on 2010 demographic data in every country. The $\delta_i$ parameters represent aging/mortality rates, which we calculated to reproduce the population age structure ($N_i$) in every country. The other parameters are listed in Table 1.

The parameters $v_i$ represent the effective vaccination rate—*i.e.*, excluding primary vaccine failures—in every age group. These parameters are connected to the age-specific effective vaccination coverage $p_{V,i}$ through the equations:

$$p_{V,i} = \frac{v_i}{v_i + \delta_i} \Rightarrow v_i = \frac{p_{V,i}}{1 - p_{V,i}}\delta_i$$

Stochastic formulation

The stochastic variant was implemented using a multinomial modification of the tau-leap algorithm (5) with a fixed time step $\Delta t = 10^{-3}$ years. At every time step, we generated multinomial samples from every source compartment to simulate all possible stochastic transitions in the model (Table S4). We then updated every compartment based on the number of entries and exits simulated for that time step. This process was iterated to simulate the model over time.

### Supplementary Figures

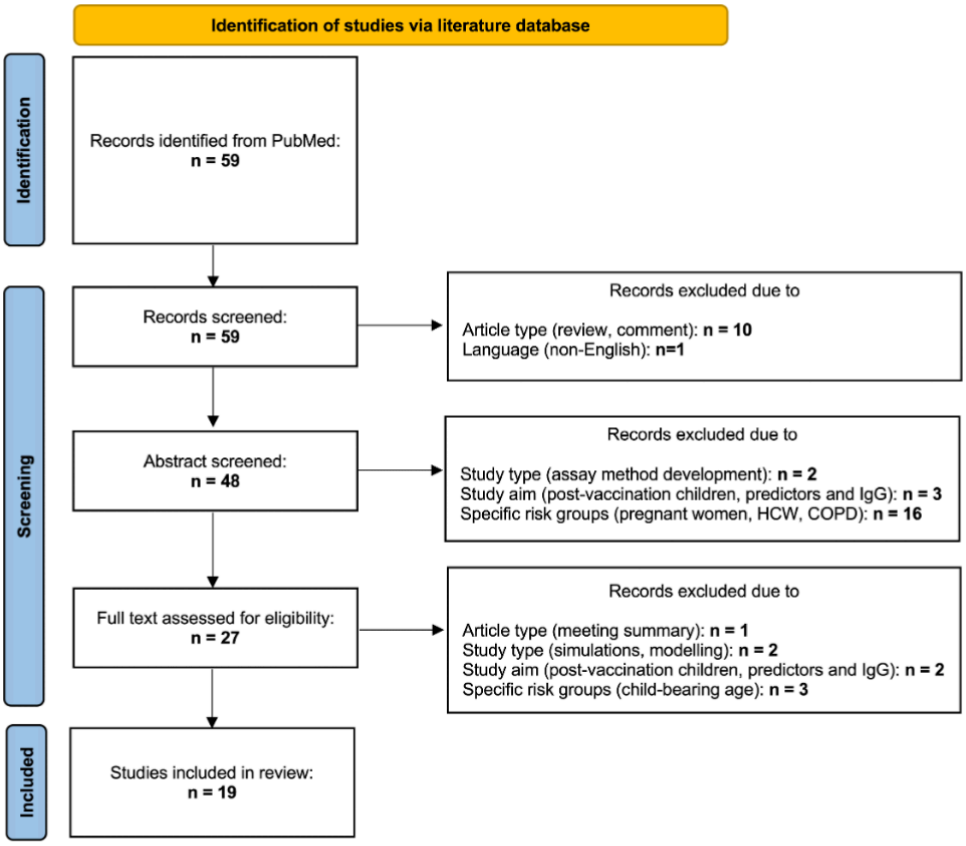


**Figure S1: PRISMA flow diagram** (6) **of the systematic review of pertussis seroprevalence studies.**

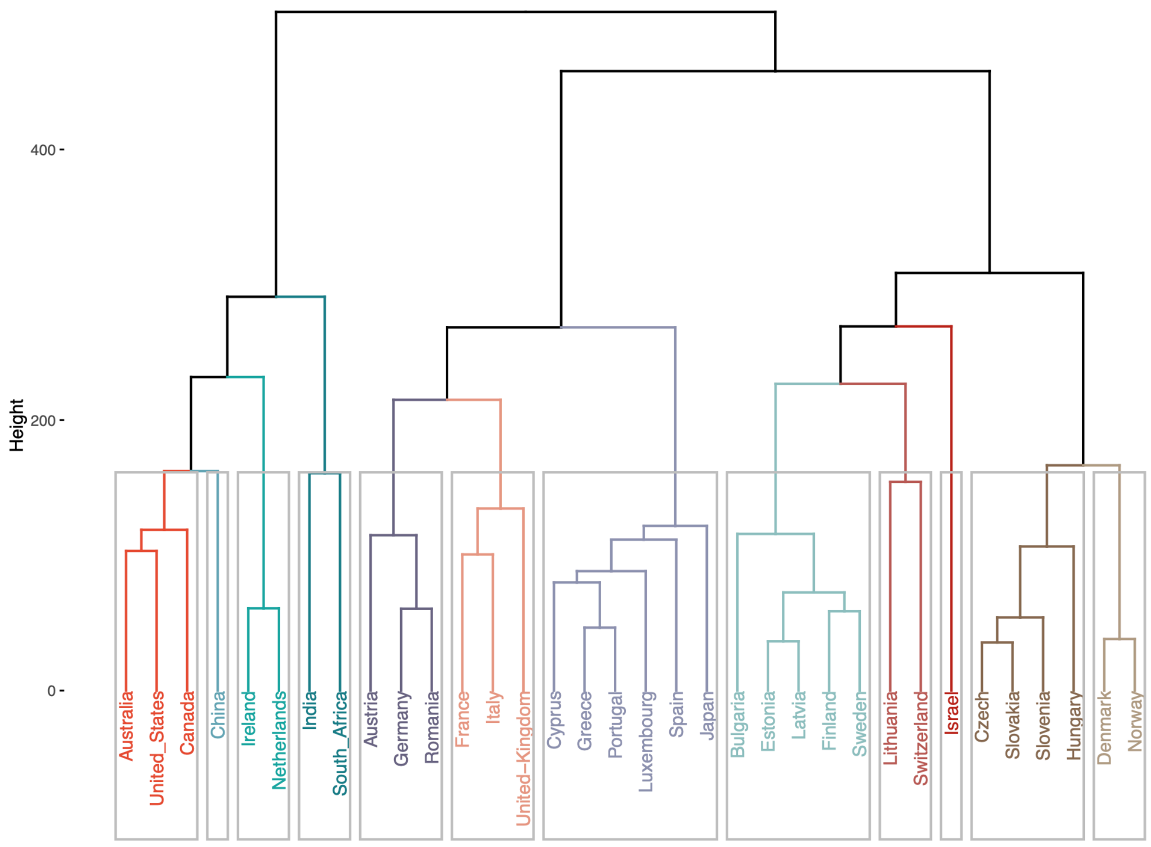


**Figure S2: Dendogram from NGM clustering**.

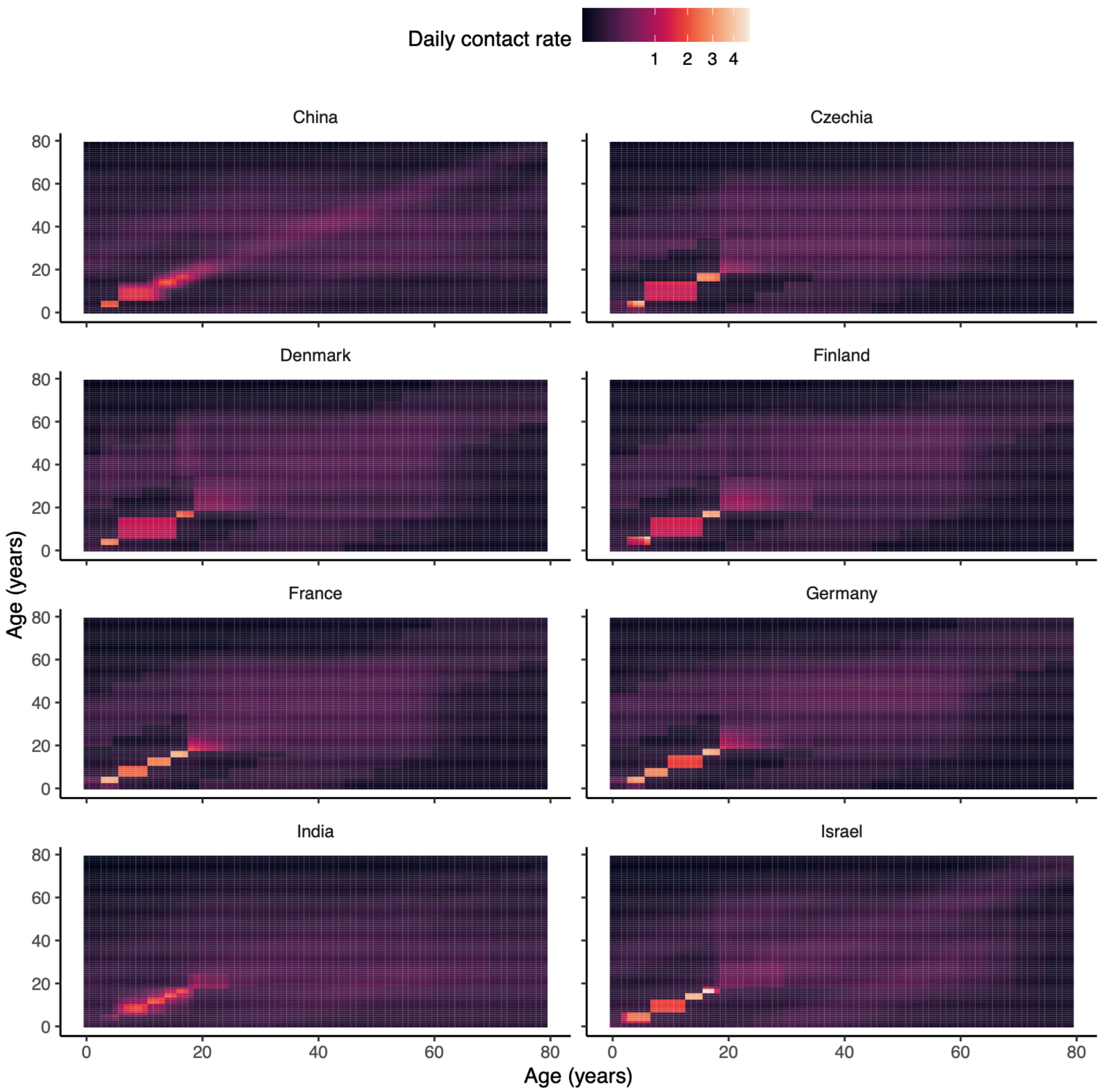


**Figure S3: Heatmaps of social contact matrices (*M*) for each country**. The color scale is transformed using a square-root function to enhance the visualization of variations at lower contact rates.

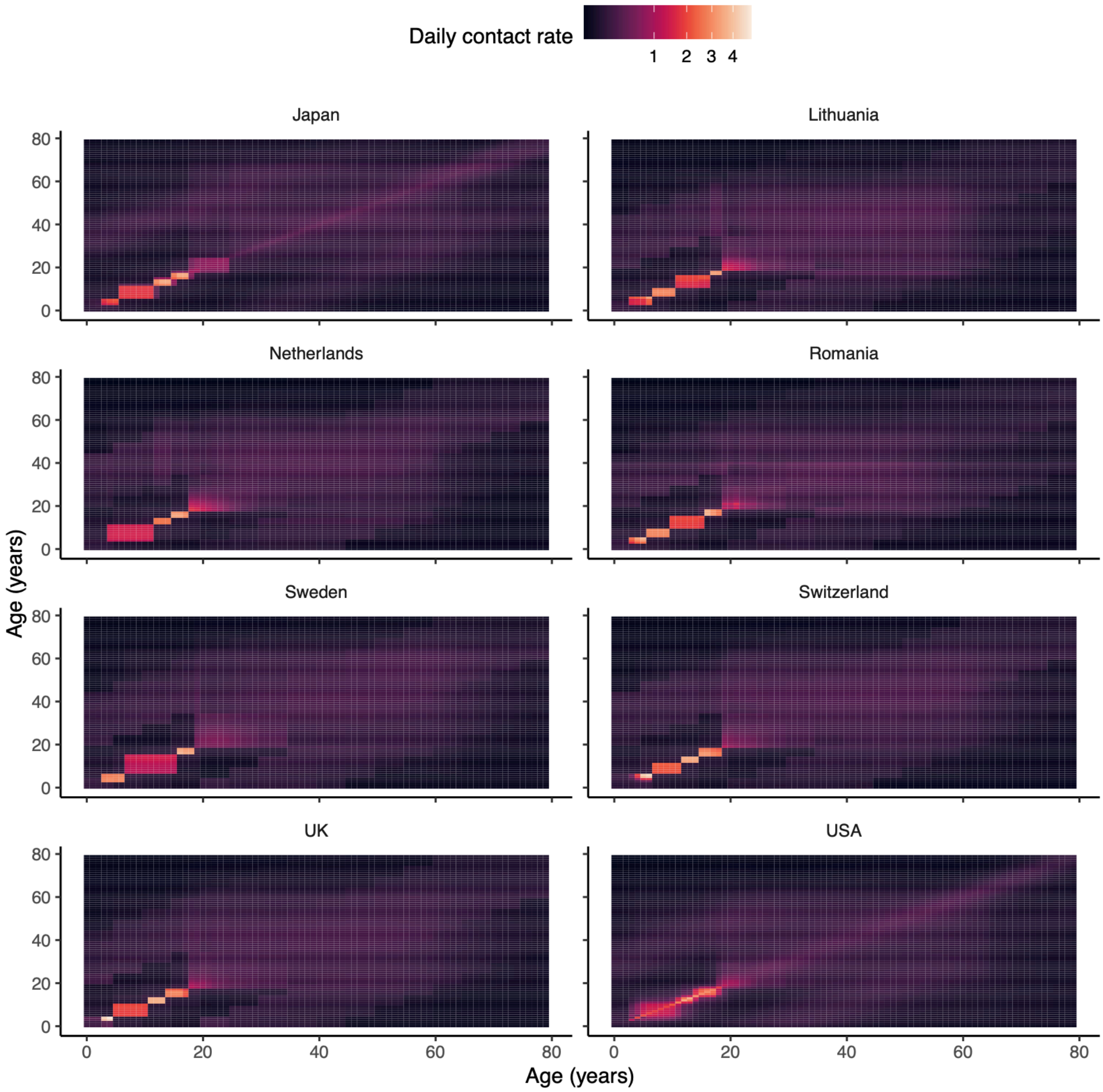


**Figure S3 (continued)**.

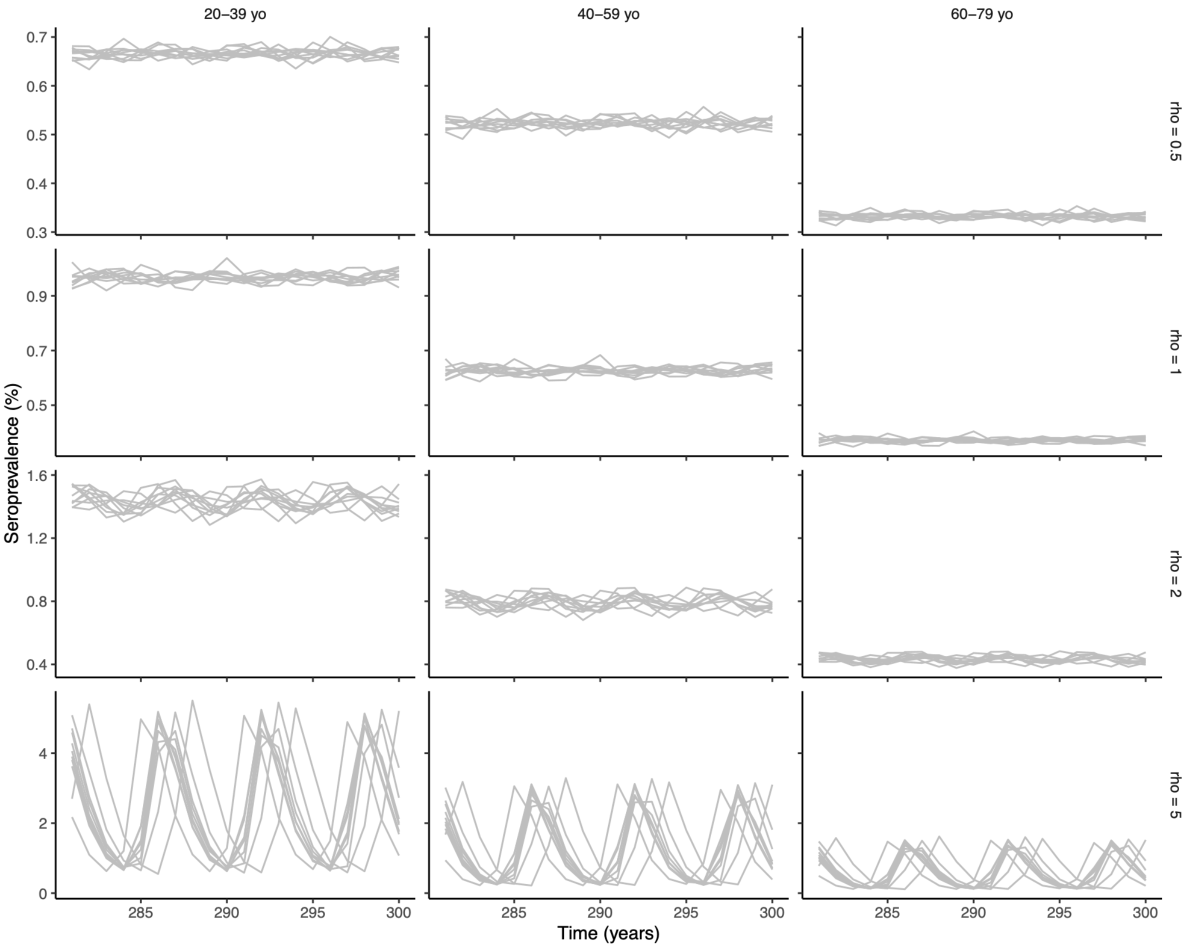


**Figure S4: Simulated time series of seroprevalence in the USA**.

**Supplementary Tables**

| Study | Population | Sample collection | IgG levels | Interpretation |
|---|---|---|---|---|
| He et al. 2022 (7) | All ages | 2020 | ≤5 IU/mL | Suggestive of no immunity |
| Liu et al., 2021 (8) | 1 d – 89 y | 2018–2019 | ≥40 IU/mL | Immune protection against infection |
| Liu et al., 2024 (9) | 0–3 m – 50 y | 2023 | ≥5 IU/mL | Immune protection against infection |
| Papagiannis et al., 2022 (10) | 30–80+ y | 2021–2022 | ≥50 IU/mL | Immune protection against infection |
| Pourakbari et al., 2025 (11) | <21 y | 2022–2023 | >11 U | Suggestive of immunity |
| Razafimahatratra et al., 2020 (12) | 6 m–15 y | 2016 | ≥5 IU/mL | Immune protection against infection |
| Sun et al., 2024 (13) | All ages | 2019–2022 | ≥20 IU/mL | Immune protection against infection |

**Table S1: Summary of seroprevalence studies aiming to assess immunity.**

| Study | Population | Sample collection | IgG levels | Interpretation |
|---|---|---|---|---|
| Bagordo et al., 2023 (14) | ≥6 y | 2019–2020 | ≥100 IU/mL | Recent infection (within last year) |
| Berber et al., 2021 (15) | 40–59 y | 2015–2018 | ≥100 IU/mL | Recent exposure |
| Chen et al., 2022 (16) | ≥15 y | 2018–2021 | ≥100 IU/mL | Recent infection (within 58.6 days) |
| | | | ≥40 IU/mL | Distant infection (within a few years) |
| Echaniz-Aviles et al., 2021 (17) | 10–25 y | 2012 | ≥94 IU/mL | Recent infection (within last year) |
| | | | ≥49 IU/mL | Distant infection (within a few years) |
| Hartanti et al., 2024 (18) | 1–14 y | 2013–2018 | ≥100 IU/mL | In age 1–4y: can indicate immune memory (vaccination) or infection<br><br>In age >4 y: high likelihood of infection |
| He et al. 2022 (7) | All ages | 2020 | ≥80 IU/mL | Recent infection in the absence of a vaccination within last year |
| Kleine et al. 2020 (19) | 1–5 y<br>6–19 y | 2013<br>2011–2012 | ≥100 IU/mL | Acute infection or recent vaccination |
| | | | [40, 100) IU/mL | Recent infection or vaccination (within last year) |
| | | | [5, 40) IU/mL | Distant infection or vaccination (more than a year ago) |
| Liu et al., 2021 (8) | 1 d–89 y | 2018–2019 | ≥100 IU/mL | Recent infection |
| | | | [40, 100) IU/mL | Possible course of pertussis |

| | | | | |
|---|---|---|---|---|
| Liu et al., 2024 (9) | 0–3 m–50 y | 2023 | ≥100 IU/mL | Acute infection |
| | | | ≥40 IU/mL | Recent infection (within last year) |
| Paradowska-Stankiewicz et al., 2021 (20) | 5–15 y | 2022–2023 | IgG ≥100 IU/mL<br>OR<br>IgG≥40 & IgA≥12 IU/mL | Recent infection |
| Razafimahatratra et al., 2020 (12) | 6 m–15 y | 2016 | ≥100 IU/mL | Acute infection |
| | | | [40, 100) IU/mL | Recent exposure |
| | | | [5, 40) IU/mL | Distant exposure |
| Silva et al., 2024 (21) | ≥4 y | | >120 IU/mL | Recent exposure |
| | | | [40, 120) IU/mL | Exposure or vaccination (within last year) |
| Sun et al., 2024 (13) | All ages | 2019–2022 | ≥80 IU/mL | Recent infection |
| Versteegen et al., 2021 (22) | All ages | 2016–2017 | >100 IU/mL | Recent infection in the absence of a vaccination in the last few years |
| Wanlapakorn et al. 2024 (23) | All ages | 2022–2023 | ≥100 IU/mL | Acute infection or recent vaccination |
| | | | [40, 100) IU/mL | Probable past exposure |
| | | | [5, 40) IU/mL | No evidence of recent exposure |
| Wehlin et al., 2021 (24) | 20–39 y | 2010–2013 | ≥100 IU/mL | Recent infection (within last year) |
| | | | [50, 100) IU/mL | Distant infection (within a few years) |

| | | | | |
|---|---|---|---|---|
| Zhang et al., 2024 (25) | All ages | 2015–2018 | IgG ≥100 IU/mL without recent vaccination<br>OR<br>IgG≥40 & IgA≥12 IU/mL | Probable recent pertussis infection |

**Table S2: Summary of seroprevalence studies aiming to assess recent exposure or infection.**

| Variable | Meaning |
| --- | --- |
| $S_1$ | Susceptible to primary infection |
| $E_1$ | Exposed, primary infection |
| $I_1$ | Infected, primary infection |
| $S_2$ | Susceptible to secondary infection |
| $E_2$ | Exposed, secondary infection |
| $I_2$ | Infected, secondary infection |
| $R$ | Recovered |
| $R_E$ | Exposed from recovered state |
| $R_{P,1}$ | Seropositive, after infection |
| $R_{P,2}$ | Seropositive, after immune boost from recovered state |
| $V$ | Vaccinated |
| $V_E$ | Exposed from vaccinated state |
| $V_P$ | Seropositive, after immune boost from vaccinated state |

**Table S3: List of model variables.**

| Source compartment | Number of exits | Transition | Rate |
|---|---|---|---|
| $S_1^{(i)}$ | 3 | $S_1^{(i)} \rightarrow S_1^{(i+1)}$ | $\delta_i$ |
| | | $S_1^{(i)} \rightarrow E_1^{(i)}$ | $\lambda_i$ |
| | | $S_1^{(i)} \rightarrow V^{(i)}$ | $v_i$ |
| $E_1^{(i)}$ | 2 | $E_1^{(i)} \rightarrow E_1^{(i+1)}$ | $\delta_i$ |
| | | $E_1^{(i)} \rightarrow I_1^{(i)}$ | $\sigma$ |
| $I_1^{(i)}$ | 2 | $I_1^{(i)} \rightarrow I_1^{(i+1)}$ | $\delta_i$ |
| | | $I_1^{(i)} \rightarrow R_{P,1}^{(i)}$ | $\gamma$ |
| $S_2^{(i)}$ | 3 | $S_2^{(i)} \rightarrow S_2^{(i+1)}$ | $\delta_i$ |
| | | $S_2^{(i)} \rightarrow E_2^{(i)}$ | $\lambda_i$ |
| | | $S_2^{(i)} \rightarrow V^{(i)}$ | $v_i$ |
| $E_2^{(i)}$ | 2 | $E_2^{(i)} \rightarrow E_2^{(i+1)}$ | $\delta_i$ |
| | | $E_2^{(i)} \rightarrow I_2^{(i)}$ | $\sigma$ |
| $I_2^{(i)}$ | 2 | $I_2^{(i)} \rightarrow I_2^{(i+1)}$ | $\delta_i$ |
| | | $I_2^{(i)} \rightarrow R_{P,1}^{(i)}$ | $\gamma$ |
| $R^{(i)}$ | 3 | $R^{(i)} \rightarrow R^{(i+1)}$ | $\delta_i$ |
| | | $R^{(i)} \rightarrow S_2^{(i)}$ | $\alpha_R$ |
| | | $R^{(i)} \rightarrow R_E^{(i)}$ | $\rho_R \lambda_i$ |
| $R_E^{(i)}$ | 2 | $R_E^{(i)} \rightarrow R_E^{(i+1)}$ | $\delta_i$ |
| | | $R_E^{(i)} \rightarrow R_{P,2}^{(i)}$ | $1/t_p$ |

| | | | |
|---|---|---|---|
| $R_{P,1}^{(i)}$ | 2 | $R_{P,1}^{(i)} \rightarrow R_{P,1}^{(i+1)}$ | $\delta_i$ |
| | | $R_{P,1}^{(i)} \rightarrow R^{(i)}$ | $1/t_n$ |
| $R_{P,2}^{(i)}$ | 2 | $R_{P,2}^{(i)} \rightarrow R_{P,2}^{(i+1)}$ | $\delta_i$ |
| | | $R_{P,2}^{(i)} \rightarrow R^{(i)}$ | $1/t_n$ |
| $V^{(i)}$ | 3 | $V^{(i)} \rightarrow V^{(i+1)}$ | $\delta_i$ |
| | | $V^{(i)} \rightarrow S_2^{(i)}$ | $\alpha_V$ |
| | | $V^{(i)} \rightarrow V_E^{(i)}$ | $\rho_V \lambda_i$ |
| $V_E^{(i)}$ | 2 | $V_E^{(i)} \rightarrow V_E^{(i+1)}$ | $\delta_i$ |
| | | $V_E^{(i)} \rightarrow V_P^{(i)}$ | $1/t_p$ |
| $V_P^{(i)}$ | 2 | $V_P^{(i)} \rightarrow V_P^{(i+1)}$ | $\delta_i$ |
| | | $V_P^{(i)} \rightarrow V^{(i)}$ | $1/t_n$ |

**Table S4: List of transitions for implementing the stochastic model variant.**

| Serosurvey | Country | Year(s) of serosurvey | Reason for exclusion |
|---|---|---|---|
| Pebody *et al.* (26) | England & Wales | 1996 | Low vaccine coverage during 1970–1989 (26) |
| | Italy | 1996 | Low vaccine coverage during 1970–1999 (26) |
| | West Germany | 1995 | Low vaccine coverage until the 1990s (26) |
| Wehlin *et al.* (24) | Belgium | 2012–13 | No SCM data from Mistry *et al.* (4) |
| | Malta | 2012 | No SCM data from Mistry *et al.* (4) |
| | Poland | 2010–11 | No SCM data from Mistry *et al.* (4) |
| | Denmark | 2012–13 | Switch to aP in 1997 (27) |
| | Finland | 2011–12 | Switch to aP in 2005 (27) |
| | Greece | 2012 | Switch to aP in 1997 (28) |
| | Hungary | 2012 | Switch to aP in 2006 (29) |
| | Italy | 2012–13 | Switch to aP in 1995 (27) |
| | Norway | 2011–13 | Switch to aP in 1998 (27) |
| | Portugal | 2012 | Switch to aP in 2006 (30) |
| | Spain | 2011 | Switch to aP in 2005 (27) |
| | Sweden | 2012 | Switch to aP in 1996 (27) |

**Table S5: Countries excluded from the analysis of seroprevalence data.**

| Country | Age of first dose | Vaccine coverage* | Age(s) of booster doses | wP start year | Year of serosurvey | Simulated period (years after wP start)** | Sources |
|---|---|---|---|---|---|---|---|
| Finland | 3 mo | ~99% | 2 yr | 1952 | 1996 | 44±5=39–49 | (26, 27, 31) |
| France | 2 mo | ~85–95% | 1 yr | 1959 | 1998 | 39±5=34–44 | (26, 27, 32) |
| East Germany | 3 mo | 85–~95% | 3 yr | 1964 | 1995 | 31±5=26–36 | (26, 33) |
| Lithuania | 3 mo | 93–97% | 1 yr | 1961 | 2012 | 38±5=33–43 | (29) |
| Netherlands | 3 mo | 96–97% | 1 yr | 1957 | 1995 | 51±5=46–56 | (26, 27) |
| Romania | 2 mo | 95–99% | 1 yr, 3 yr | 1961 | 2010–11 | 50±5=45–55 | (29) |

**Table S6: Characteristics of countries included in the analysis of seroprevalence data.** *Data represent the coverage with four doses in Lithuania and Romania (source: (29)), with three doses in the other countries (source: (26)). Based on these data, we assumed, for simplicity, an effective vaccine coverage of 90% for all doses, corresponding to a 95% vaccine coverage and a 5% probability of primary vaccine failure (2). **We defined a simulated period of ±5 years after wP start to take into account potential uncertainties in the start date and initial coverage of wP programs.
